# Elucidating mechanisms of genetic cross-disease associations: an integrative approach implicates protein C as a causal pathway in arterial and venous diseases

**DOI:** 10.1101/2020.03.16.20036822

**Authors:** David Stacey, Lingyan Chen, Joanna M. M. Howson, Amy M. Mason, Stephen Burgess, Stephen MacDonald, Jonathan Langdown, Harriett McKinney, Kate Downes, Neda Farahi, James E. Peters, Saonli Basu, James S. Pankow, Nathan Pankratz, Weihong Tang, Maria Sabater-Lleal, Paul S. de Vries, Nicholas L. Smith, CHARGE Hemostasis Working Group, Amy D. Gelinas, Daniel J. Schneider, Nebojsa Janjic, Charlotte Summers, Edwin R. Chilvers, John Danesh, Dirk S. Paul

## Abstract

Genome-wide association studies have identified many individual genetic loci associated with multiple complex traits and common diseases. There are, however, few examples where the molecular basis of such pleiotropy has been elucidated. To address this challenge, we describe an integrative approach, focusing on the p.Ser219Gly (rs867186 A>G) variant in the *PROCR* gene (encoding the endothelial protein C receptor, EPCR), which has been associated with lower coronary artery disease (CAD) risk but higher venous thromboembolism (VTE) risk. In a phenome scan of 12 cardiometabolic diseases and 24 molecular factors, we found that *PROCR*-219Gly associated with higher plasma levels of zymogenic and activated protein C as well as coagulation factor VII. Using statistical colocalization and Mendelian randomization analyses, we uncovered shared genetic etiology across activated protein C, factor VII, CAD and VTE, identifying p.S219G as the likely causal variant at the locus. In a recall-by-genotype study of 52 healthy volunteers stratified by p.S219G, we detected 2.5-fold higher soluble EPCR levels and 1.2-fold higher protein C levels in plasma per effect allele, suggesting the allele induces EPCR shedding from the membrane of endothelial cells. Finally, in cell adhesion assays, we found that increasing concentrations of activated protein C, but not soluble EPCR, reduced leukocyte–endothelial cell adhesion, a marker for vascular inflammation. These results support a role for protein C as a causal factor in arterial and venous diseases, suggesting that *PROCR*-219Gly protects against CAD through anti-inflammatory mechanisms while it promotes VTE risk through pro-thrombotic mechanisms. Overall, our study illustrates a multi-modal approach that can help reveal molecular underpinnings of cross-disease associations.

## Introduction

Genome-wide association studies (GWAS) have revealed widespread pleiotropy of disease-associated genetic variants. A recent study of cross-phenotype genetic data in the UK Biobank has shown that 96% of trait-associated variants (minor allele frequency (MAF) ≥1%) are associated with more than one ICD-10 code, with some showing associations with more than 50 codes ^1^. The vast majority of these pleiotropic variants were found to impact the risk of multiple diseases in a directionally consistent manner, but 1.9% of loci (excluding the major histocompatibility complex) showed evidence of both higher and lower risk effects attributable to the same allele ^1^. One such example is rs9349379 A>G, a well-characterized regulatory variant at the *PHACTR1*-*EDN1* locus, which is associated with a *higher* risk of coronary artery disease but a *lower* risk of four other vascular diseases including migraine headache and hypertension ^2^.

Another example of a pleiotropic variant is p.Ser219Gly (rs867186 A>G) in the *PROCR* gene, which encodes the endothelial protein C receptor (EPCR), a key regulator of the protein C (PC) pathway. The minor G allele of this variant has been shown to correlate with a *lower* risk of CAD ^3,4^ and myocardial infarction ^5^, but a *higher* risk of venous thromboembolism (VTE) ^6-8^. This pattern of opposing associations seems paradoxical because several conventional cardiovascular risk factors (e.g. measures of adiposity) show directionally concordant associations for CAD and VTE ^9^. Further, GWAS of cardiovascular intermediate traits have reported tentative associations between rs867186-G and components of the coagulation cascade, including higher plasma levels of PC ^10^ and coagulation factor VII ^11,12^, as well as shorter prothrombin time ^13^. However, the causal relevance of these intermediate traits to cardiovascular diseases remains uncertain.

The thrombomodulin–protein C pathway serves as a key mediator of the cross-talk between coagulation and inflammatory processes. It comprises molecular components that can respond to a range of pathophysiological environments in different vascular beds ^14-16^. At the vascular endothelium, thrombomodulin binds to thrombin, directly inhibiting its clotting and cell activation potential and converting PC to APC (reviewed in ^16,17^). The activation of PC by the thrombin–thrombomodulin complex is markedly enhanced when PC is presented by EPCR ^18^, a type I transmembrane protein that is mainly expressed on the endothelium of large blood vessels. ^19,20^ Once APC dissociates from EPCR, it binds to protein S to inactivate the coagulation factors Va and VIIIa, thereby inhibiting further thrombin generation. In addition, APC promotes fibrinolysis by decreasing the levels of plasminogen-activator inhibitor type 1 (PAI-1), and reduces inflammation by inhibiting the production of tumor necrosis factor (TNF)-α and interleukin(IL)-1β (reviewed in ^16,17^).

A soluble form of EPCR (sEPCR) is present in plasma, which is generated by ectodomain shedding of EPCR from the endothelium. Plasma sEPCR levels in healthy individuals display a bimodal distribution, with higher levels being associated with one of the four frequent haplotypes constituting *PROCR* ^8,21-24^. This haplotype (denoted A3 or H3) is tagged by the minor allele of the p.S219G variant. Functional studies showed that the variant results in increased shedding of EPCR from the endothelial surface by rendering the receptor more sensitive to cleavage by metalloprotease ^22^ and by forming an alternatively spliced, truncated transcript ^25^. The shedding is effectively regulated by TNF-α and IL-1β ^26^. sEPCR retains its ability to bind both PC and APC but does not enhance PC activation ^27,28^. However, the precise molecular mechanism underlying the *PROCR*-p.S219G functional variant and its influence on the cardiovascular intermediate phenotypes that may mediate the risk of CAD and VTE is incompletely understood.

In this study, we aim (1) to systematically assess the association of the *PROCR*-p.S219G pleiotropic variant with a range of cardiometabolic outcomes and relevant risk factors; (2) to evaluate causality of individual components of the protein C pathway on cardiovascular diseases; and (3) to help uncover the molecular and cellular chain-of-events that connect the *PROCR*-219Gly allele to a lower risk of CAD but a higher risk of VTE. The results of our integrative epidemiological and functional analyses (**Fig. 1**) reveal new insights underlying the *PROCR* association locus for arterial and venous diseases and have potential implications for the development of therapeutic strategies targeting components of the protein C pathway.

**Figure 1.**
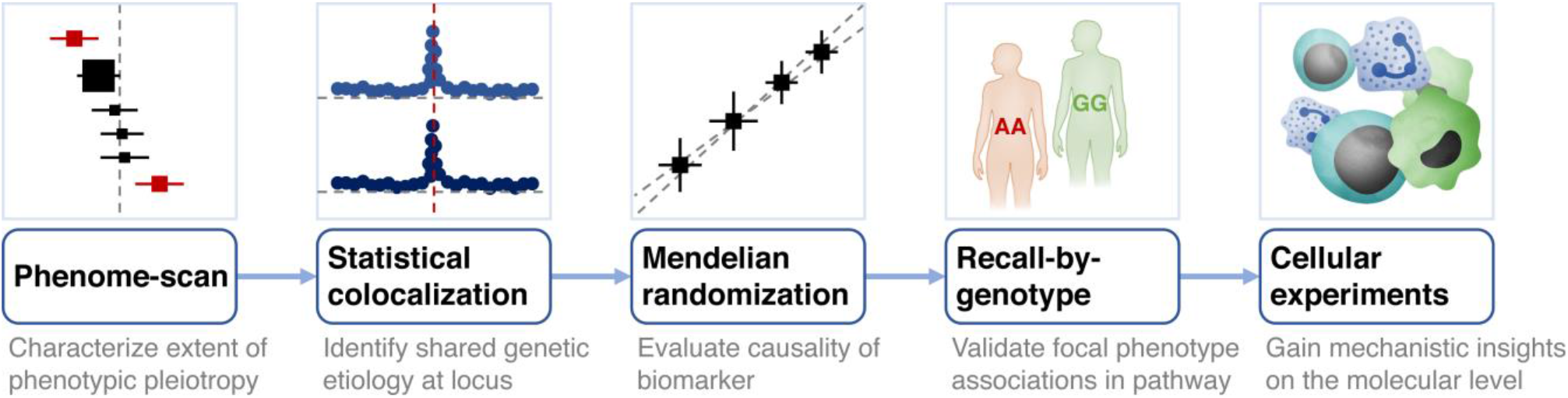
Schematic overview of the study design to elucidate molecular underpinnings of cross-disease associations. <u>Credits:</u> The immune response, Big Picture (http://bigpictureeducation.com/).

## Results

### Association of PROCR-p.S219G with cardiovascular diseases and risk factors

To search for associations of *PROCR*-p.S219G with multiple cardiovascular and related health outcomes, we conducted a phenome-wide association analysis across 1,402 broad electronic health record-derived ICD-codes from the UK Biobank. The association of *PROCR*-p.S219G with each of these codes was tested using SAIGE ^29^, a generalized mixed model association test that accounts for case-control imbalance and sample relatedness (**Methods**). The data implicated diseases of the circulatory system, e.g. phlebitis/thrombophlebitis (PheWAS code 451; *P*=4.2×10^−8^) and coronary atherosclerosis (PheWAS code 411.4; *P*=2.9×10^−5^) (**Fig. 2A**).

**Figure 2.**
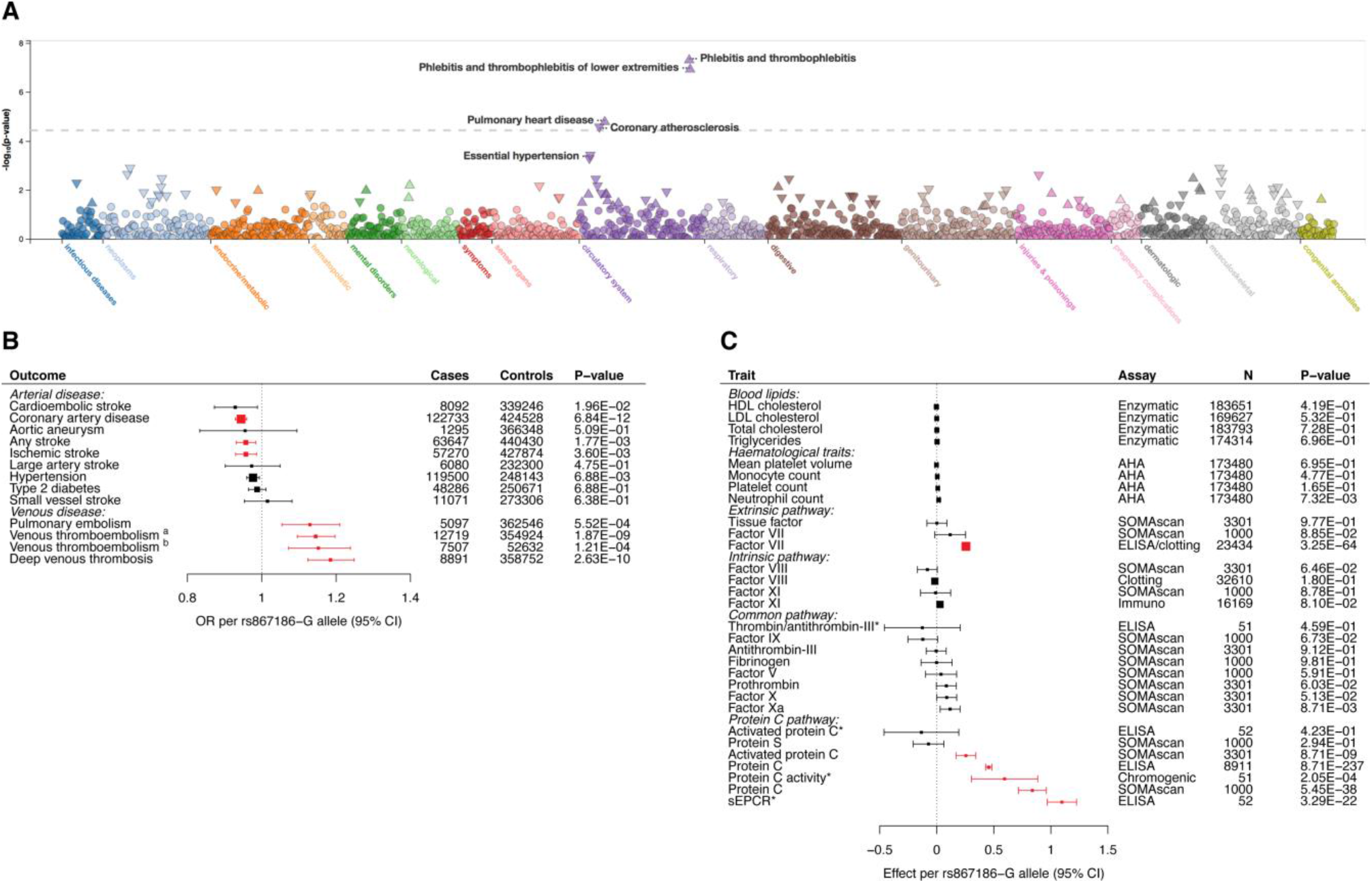
Association of PROCR-219Gly with a range of health outcomes and circulating cardiovascular biomarkers. **(A)** Phenome-wide association scan of PROCR-p.S219G (rs867186) across 1,402 broad electronic health record-derived ICD-codes from the UK Biobank. The data were extracted from PheWeb v1.1.17 (http://pheweb.sph.umich.edu/SAIGE-UKB/variant/20:33764554-A-G). **(B)** The forest plot shows the associations of the minor (G) allele of rs867186 genotype with different cardiovascular conditions. The following association statistics were retrieved: stroke outcomes from the MEGASTROKE consortium ^37^; venous thromboembolism outcomes from the INVENT consortium (a) ^7^ or UK Biobank (b); hypertension and aortic aneurysm from UK Biobank; coronary artery disease from van der Harst et al. ^4^; and type 2 diabetes from Mahajan et al ^38^. Associations that passed correction for multiple testing in this analysis (P=0.05/13 traits=3.85×10^−3^) are highlighted in red. **Suppl. Table 1** provides the association statistics for all traits, as well as data sources and references. **(C)** The forest plot shows the associations of the minor (G) allele of rs867186 (PROCR-219Gly) with clinical biomarkers (blood lipids, hematological traits) and plasma proteins of the coagulation cascade (extrinsic, intrinsic and common pathways) and protein C pathways. To enable a comparison of the magnitude of the associations across several different traits, we conducted analyses with standardized units of measurement for each trait. Associations are presented as per-allele changes in the traits expressed as standard deviations. The following association statistics were retrieved: blood lipids from the Global Lipids Genetics consortium ^39^; hematological traits from Astle et al. ^40^; and plasma proteins of the coagulation cascade and protein C pathway from the ARIC study ^10^, CHARGE consortium ^35,41,42^, Sun et al. ^31^, Suhre et al. ^43^ and de novo measurements obtained from the recall-by-genotype study described in this report (indicated with an asterisk). The type of assay for each measurement is indicated. Associations that passed correction for multiple testing in this analysis (P=0.05/30 traits=1.67×10^−3^) are highlighted in red. **Suppl. Table 1** provides the association statistics for all traits, as well as data sources and references. Box sizes are proportional to inverse-variance weights. Error bars show the 95% confidence intervals. <u>Abbreviations:</u> AHA, automated hematology analyzer; ELISA, enzyme-linked immunosorbent assay.

Next, we performed a more focused phenome scan of the circulatory system. For each trait, we retrieved the largest available genetic association dataset (**Methods**; **Suppl. Table 1**). We found that the minor (G) allele of rs867186 (219Gly) was consistently associated with a higher risk of VTE in the UK Biobank (odds ratio (OR)=1.15 [95% confidence interval (CI)=1.10, 1.20]; *P*=1.87×10^−9^) and INVENT (OR=1.15 [1.07, 1.24]; *P*=1.21×10^−4^) studies (**Fig. 2B**). We also observed a higher risk of deep vein thrombosis (DVT; OR=1.18 [1.12, 1.25]; *P*=2.63×10^−10^) and pulmonary embolism (OR=1.13 [1.05, 1.21]; *P*=5.52×10^−4^) in UK Biobank, with both of these conditions being manifestations of VTE (**Fig. 2B**). In contrast, rs867186-G was associated with a lower risk of CAD in a large GWAS meta-analysis of the UK Biobank and CARDIoGRAMplusC4D consortium (OR=0.94 [0.93, 0.96]; *P*=6.84×10^−12^) (**Fig. 2B**). Further, we detected a tentative association of rs867186-G with a lower risk of any stroke (OR=0.96 [0.93, 0.98]; *P*=1.77×10^−3^) and ischemic stroke (OR=0.96 [0.93, 0.99]; *P*=3.60×10^−3^) in the MEGASTROKE consortium (**Fig. 2B**). Collectively, these data suggest that individuals carrying rs867186-G alleles have lower susceptibility to arterial thrombotic diseases but a higher risk of venous diseases.

To explore the molecular basis for this association pattern, we associated the rs867186-G allele with various intermediate traits related to the cardiovascular system (**Methods**). In particular, we focused on traits that directly influence the protein C pathway. We found that rs867186-G correlates strongly with higher PC levels in plasma, measured using an enzyme-linked immunosorbent assay (ELISA) in the ARIC study (per-allele effect=0.46 standard deviation (SD) [0.43, 0.48]; *P*=8.71×10^−237^) (**Fig. 2C**). The allelic effect was also observed using the highly sensitive, multiplexed SomaScan assay in the KORA study (0.84 SD [0.72, 0.96]; *P*=5.45×10^−38^). This assay quantifies the relative concentrations of plasma proteins or protein complexes using modified aptamers (‘SOMAmer reagents’) ^30,31^. Further, the allele was significantly associated with elevated plasma levels of APC (0.26 SD [0.17, 0.34]; *P*=8.71×10^−9^) and coagulation factor VII (0.26 SD [0.25, 0.26]; *P*=3.25×10^−64^) (**Fig. 2C**). We neither detected associations with plasma levels of other measured proteins in the coagulation cascade and protein C pathway, including protein S (the cofactor of APC), nor with risk factors for thrombosis, including fibrinogen, von Willebrand factor (vWF), plasminogen activator inhibitor-1 (PAI-1) and the thrombolytic agent tissue plasminogen activator (tPA) (*P*>0.05) ^32-35^ (**Fig. 2C**). However, we did observe a suggestive signal for D-dimer, a fibrin degradation product, with higher levels observed in G-allele carriers (per-allele effect=0.05 log(ng/dL); *P*=3.70×10^−6^) ^36^. Finally, rs867186-G was not associated with conventional cardiovascular risk factors, including lipid levels, type 2 diabetes and hypertension (**Figs. 2B, C**).

We investigated a subset of the molecular intermediate traits, including PC, APC and FVII, using the SomaScan assay. To confirm the specificity of the binding events, we measured the binding activity of the PC and APC SOMAmer reagents to a range of relevant proteins, specifically, PC, APC, sEPCR, thrombin, FV, FVIIa, Protein S and thrombomodulin (**Methods**). We confirmed that the APC SOMAmers bind the protein in a specific manner. However, we found that the PC SOMAmer binds to both the zymogenic and activated form of protein C (**Suppl. Table 2**), which may contribute to the observed difference in the magnitude of effect sizes observed for the immuno- and SomaScan assays (**Fig. 2C**). Additionally, we confirmed that the presence of relevant binding partners of PC and APC do not interfere with SOMAmer binding (**Suppl. Table 2**).

### Identification of shared genetic etiology at the PROCR locus

Despite data showing associations of the rs867186 variant at the *PROCR* locus with CAD and VTE, it has been uncertain whether they reflect a shared causal variant and mechanism. To formally test this hypothesis, we performed statistical colocalization analyses. We applied a Bayesian algorithm, Hypothesis Prioritization in multi-trait Colocalization (HyPrColoc) ^44^, which allows for the assessment of colocalization across multiple complex traits simultaneously (**Methods**). We found colocalization of the genetic association data of CAD and VTE as well as factor VII, PC and APC levels at the *PROCR* locus, with a posterior probability of colocalization of 99.37% (**Fig. 3A**). The variant rs867186 was found to be the likely causal variant at the locus explaining 99.31% of the posterior probability (**Fig. 3B**). Thus, these data support the hypothesis of a common genetic mechanism underlying the *PROCR* locus.

**Figure 3.**
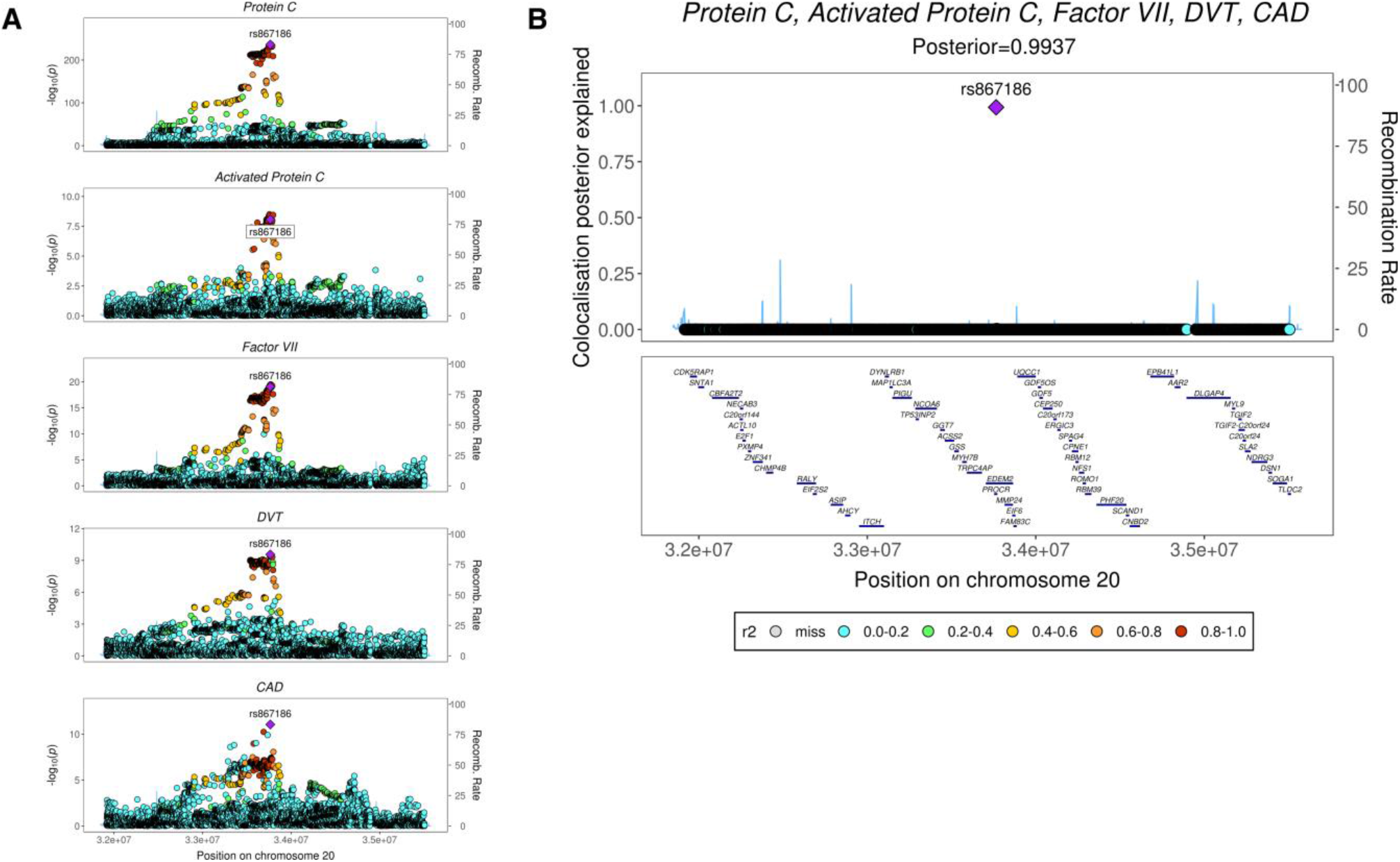
Statistical colocalization of cardiovascular outcomes and traits at the PROCR locus. **(A)** Regional association plots at the PROCR gene locus, showing the genetic association with coagulation factor VII, protein C, activated protein C, DVT and CAD. Details about the statistical analysis and source of the data are given in the **Methods** section. Color key indicates r^2^ with the respective lead variants in the GWAS. **(B)** Plot showing the posterior probabilities of all genetic variants at the chr20q11.22 locus tested in the colocalization analysis.

### Causal evaluation of protein C in arterial and venous diseases

The association data suggest that genetic variants at the *PROCR* locus influence PC, APC and FVII abundance and susceptibility to CAD and DVT. However, these data do not necessarily imply that these molecular traits have a causal relationship with the disease phenotypes. To help define this relationship, we conducted Mendelian randomization (MR) analyses, using genetic variants as instrumental variables to avoid confounding and reverse causation ^45^. We constructed a multi-allelic genetic score to estimate the causal associations between PC and cardiovascular outcomes (**Methods**). The score comprised of approximately independent (r^2^ <0.1) SNPs at the *PROCR* region with *P*-value ≤5×10^−8^ (**Methods**; **Suppl. Table 3**). Our data showed that every genetically-predicted increment (per 1 SD) in PC levels is associated with a lower risk of CAD (OR=0.88 [0.86, 0.90]; *P*=4.17×10^−24^) (**Fig. 4A**), any stroke, ischemic stroke and cardioembolic stroke, as well as a higher risk of VTE (OR=1.24 [1.17, 1.32]; *P*=1.05×10^−11^), DVT (OR=1.34 [1.25, 1.44]; *P*=8.70×10^−16^) (**Fig. 4B**) and pulmonary embolism (**Table 1**; **Suppl. Fig 1**). We also performed these analyses with APC, resulting in similar effect sizes and association *P*-values (**Suppl. Table 4**). Findings were robust to the use of a range of different MR approaches, i.e. inverse-variance weighting (IVW) method, MR-Egger regression and MR-PRESSO (**Methods**). We conducted further sensitivity analyses, confirming the validity of our results to potential violations of the MR assumptions (**Methods**; **Suppl. Fig. 2**). Finally, we applied reverse MR to evaluate evidence for causal effects in the reverse direction by modelling disease phenotypes as the exposure and PC level as the outcome using genome-wide significant predictors of disease (**Methods**). These analyses revealed no reverse causality of CAD or DVT/VTE on the levels of PC (**Table 1**). Taken together, these analyses provide evidence of causal relationships between the levels of zymogenic and activated protein C and CAD and VTE outcomes, in opposite directions.

**Table 1.** Mendelian randomization estimates for the effect of genetically determined levels of protein C and activated protein C on the risk of vascular diseases and traits. Full details of the results from the different MR analyses, including details of data sources and number of cases, are reported in **Suppl. Table 4**.

| Exposure | Outcome | No of SNPs <sup>1</sup> | Inverse variance weighting |  | Q statistic |  |
| --- | --- | --- | --- | --- | --- | --- |
|  |  |  | Odds ratio [95% CI] <sup>2</sup> | P-value | Heterogeneity | P-value |
| Forward MR |  |  |  |  |  |  |
| Protein C | Coronary artery disease | 19 | 0.88 [0.86, 0.90] | 4.17×10<sup>-24</sup> | 17.24 | 0.51 |
| Protein C | Deep venous thrombosis | 18 | 1.34 [1.25, 1.44] | 8.70×10<sup>-16</sup> | 14.21 | 0.65 |
| Protein C | Pulmonary embolism | 18 | 1.17 [1.06, 1.29] | 2.65×10<sup>-3</sup> | 19.01 | 0.33 |
| Protein C | Venous thromboembolism | 18 | 1.24 [1.17, 1.32] | 1.05×10<sup>-11</sup> | 17.80 | 0.40 |
| Protein C | Any stroke | 18 | 0.90 [0.86, 0.94] | 2.86×10<sup>-6</sup> | 13.79 | 0.68 |
| Protein C | Ischemic stroke | 18 | 0.90 [0.86, 0.95] | 3.77×10<sup>-5</sup> | 12.48 | 0.77 |
| Protein C | Large-artery stroke | 18 | 0.94 [0.83, 1.07] | 0.376 | 15.86 | 0.53 |
| Protein C | Cardioembolic stroke | 18 | 0.85 [0.77, 0.94] | 2.10×10<sup>-3</sup> | 17.83 | 0.40 |
| Protein C | Small-vessel stroke | 18 | 0.94 [0.82, 1.07] | 0.332 | 21.24 | 0.22 |
| Reverse MR |  |  |  |  |  |  |
| Coronary artery disease | APC | 138 | 0.99 [0.90, 1.09] | 0.837 | 144.15 | 0.32 |
| Deep venous thrombosis | APC | 17 | 0.94 [0.87, 1.01] | 0.101 | 18.23 | 0.31 |
| Venous thromboembolism | APC | 19 | 0.94 [0.86, 1.02] | 0.157 | 22.71 | 0.20 |

<sup>1</sup>Number of SNPs as instrumental variants for PC or APC. <sup>2</sup>Represents increase/decrease of risk per SD increase in PC or APC levels.

**Figure 4.**
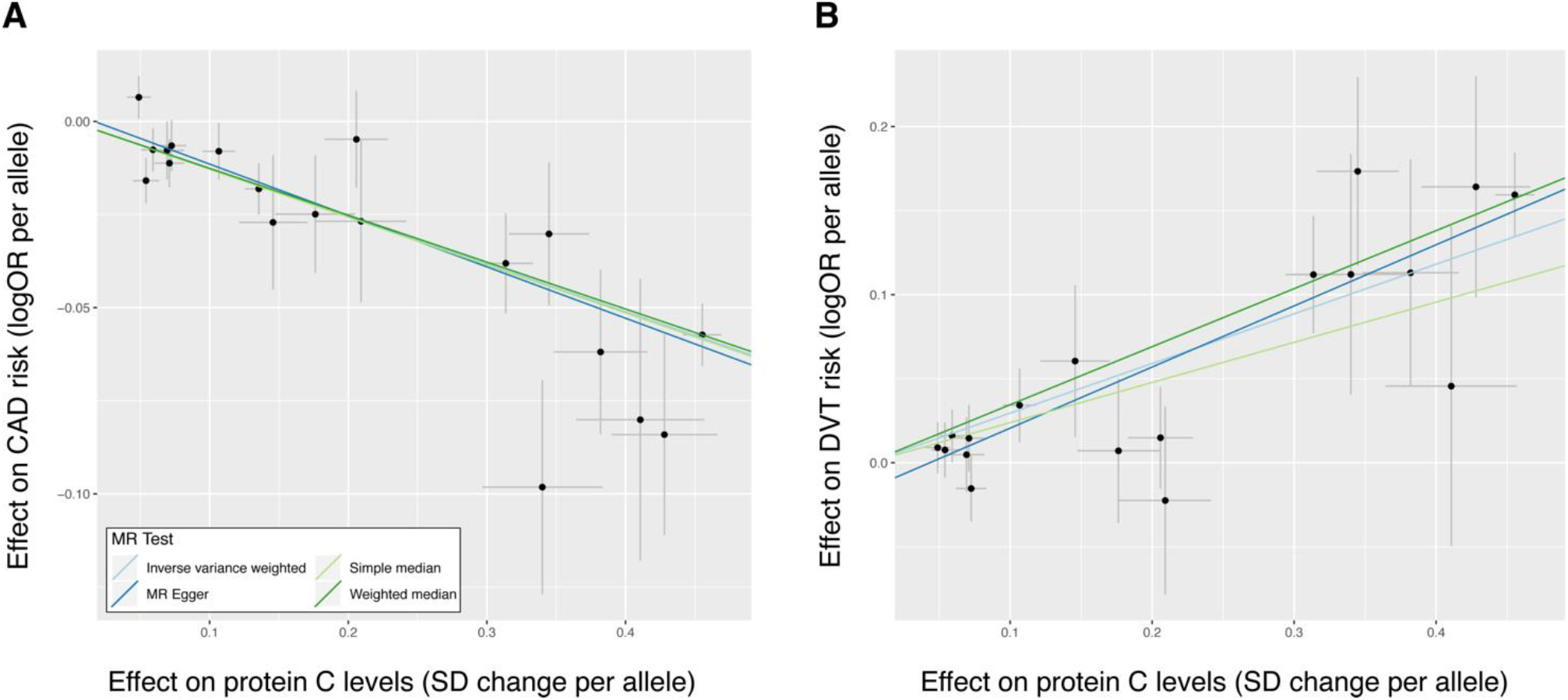
Plasma levels of protein C and risk of CAD and DVT. Scatter plot showing the estimated effect sizes and 95% confidence intervals on plasma protein C levels (from linear regression) and risk of **(A)** CAD and **(B)** DVT (from logistic regression) for each variant used in the genetic score. The different regression lines indicate the effect sizes as calculated by different MR tests (**Methods**). An overview of the results for other cardiovascular outcomes is shown in **Suppl. Fig. 1**. The corresponding sensitivity analyses are provided in **Suppl. Fig. 2**. Data sources used for these analyses are provided in **Suppl. Table 1**.

### Validation of ‘focal’ phenotype associations in the protein C pathway

To determine the molecular and cellular effects of rs867186, the causal variant at the *PROCR* locus, we performed a recall-by-genotype study. Such recall-studies allow for the strict control of experimental conditions (e.g. identical processing of blood samples), statistical efficiency (i.e. balanced recruitment based on genotype independent of MAF) and deep-phenotypic characterization of the collected samples (e.g. *in vitro* challenge experiments) (reviewed in ^46^). From a genotyped panel of healthy volunteers, we selected 52 individuals stratified by rs867186 genotype and matched for sex and age (**Methods**). In these individuals, we measured four biomarkers in plasma that represent focal phenotypes that describe the functional state of the protein C pathway, i.e. levels of protein C (inferred from a chromogenic assay measuring PC activation in response to an exogenous stimulus), APC, sEPCR and thrombin-antithrombin (TAT) complex (**Methods**). We found that the minor (G) allele of rs867186 associated with higher plasma levels of sEPCR (β=1.10, *P*=3.29×10^−22^) (**Fig. 5**). This finding is consistent with the previous literature ^8,24,47-51^ and provides further evidence that *PROCR*-219Gly leads to increased shedding of EPCR. We also found that the G allele associated with elevated PC activity, a marker for PC levels (β=0.59, *P*=2.05×10^−4^) (**Fig. 5**). These data are concordant with the data that we report from the epidemiological studies above (**Fig. 2C**), which were generated using the SomaScan assay. We did not observe genotypic effects on plasma levels of either APC (β=-0.14, *P*=0.42) or TAT complex (β=-0.13, *P*=0.46) (**Fig. 5**). These data provide a direct comparison of the genotypic effect of the *PROCR* functional variant on the functional PC pathway, showing that the strongest effects relate to PC and sEPCR.

**Figure 5.**
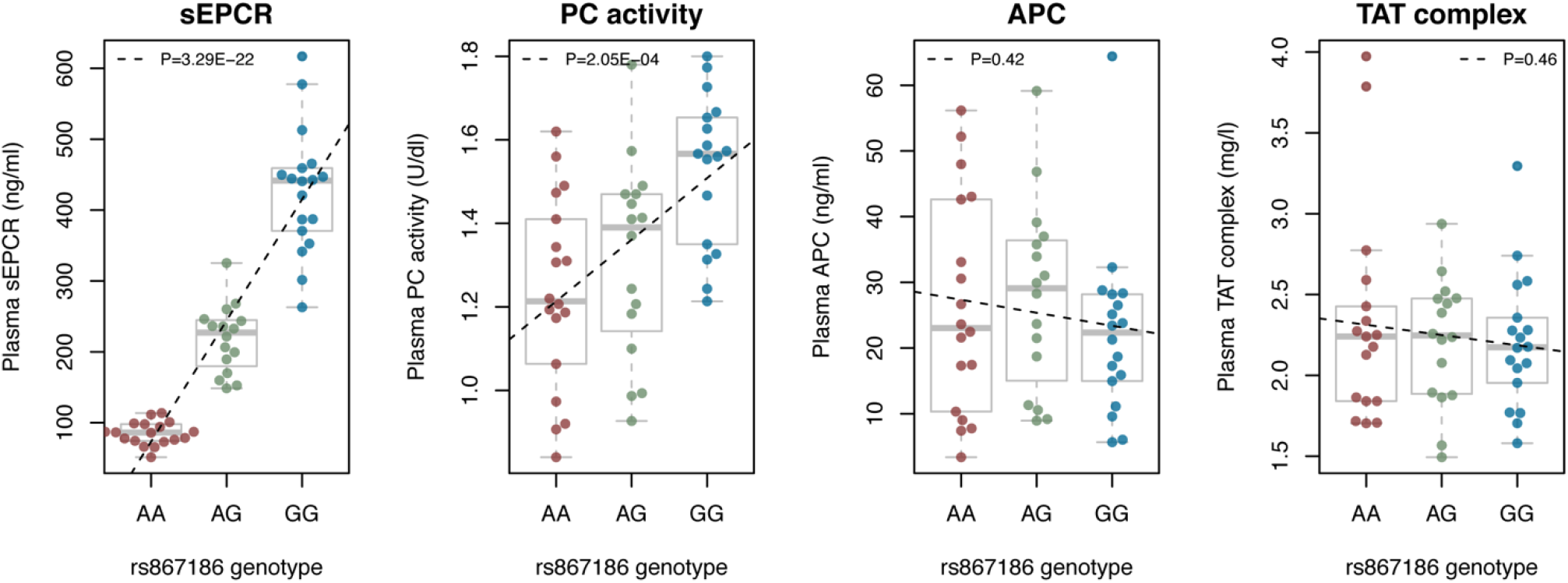
Measurement of plasma biomarkers in 52 individuals stratified by rs867186 genotype. We measured plasma levels of sEPCR, APC and TAT complex using immunoassays. PC levels were measured using a chromogenic assay. Boxplots represent the distribution of the plasma levels of each biomarker as a function of rs867186 genotype in a total of 52 individuals. Dashed lines indicate the fitted linear regression model for biomarker∼genotype. P-values (additive regression model) are indicated.

### Quantification of EPCR surface levels on leukocytes and endothelial cells

We next aimed to determine the downstream molecular consequences of elevated plasma sEPCR and PC (APC) levels due to the rs867186 genotype. Using flow cytometric analyses in the human umbilical vein endothelial cell line (HUVEC) ^52^ and human monocytic cell line U937 ^53^, we detected protein expression levels of EPCR on both HUVECs (**Suppl. Figs. 3, 4**) and U937 cells (**Suppl. Fig. 5**). The EPCR surface expression was ∼75% higher in HUVECs compared to U937 cells (**Suppl. Figs. 4, 5**). Differentiation of U937 cells into macrophages using phorbol 12-myristate 13-acetate (PMA) (**Methods**) resulted in a reduction of EPCR expression by ∼25% (**Suppl. Fig. 5**). Notably, we did not detect EPCR expression on human primary monocytes and neutrophils (**Suppl. Fig. 6**). Taken together, our data suggest that EPCR is predominantly expressed on endothelial cells, consistent with the available gene expression data generated across hematopoietic and vascular cells (**Suppl. Fig. 7**). Therefore, our data indicate that endothelial cells represent the primary source of sEPCR.

### Effect of sEPCR and APC on leukocyte–endothelial cell adhesion

Leukocyte–endothelial cell adhesion is an important step in atherosclerosis that triggers vascular infiltration of monocytes and subsequently leads to microvascular inflammation ^54^. Findings from previous *in vitro* studies have highlighted both EPCR and APC as potential candidates involved in the modulation of leukocyte–endothelial cell adhesion. For example, sEPCR is an established binding partner for the integrin macrophage-1 antigen (Mac-1) ^55^, which is expressed on the surface of activated leukocytes and is a key mediator of adhesion to the endothelium. Furthermore, it has been shown that exposure of endothelial cells to APC leads to a reduction in the surface levels of cellular adhesion molecules ^56^. Consequently, we sought to determine the effects of increasing concentrations of recombinant human sEPCR and APC on leukocyte–endothelial cell adhesion using an *in vitro* static adhesion model. In brief, U937 cells were differentiated into macrophage-like cells using PMA and then dispensed onto a monolayer of TNF-α-activated HUVECs (**Methods**). Cell adhesion events were quantified following incubation with increasing concentrations of anti-Mac-1 antibody (positive control) and recombinant sEPCR and APC (**Methods**). We found that increasing concentrations of anti-Mac-1 antibody (compared to an IgG control; *P*=0.029) and APC (*P*=0.048) but not sEPCR (*P*=0.789) lead to a reduction of adhesion events (**Fig. 6**). Together, these data suggest that in carriers of the *PROCR*-219Gly genotype, who exhibit elevated APC levels, the lower genetic susceptibility to arterial disease may be due to a reduced number of leukocyte–endothelial cell adhesion events at sites of vascular inflammation.

**Figure 6.**
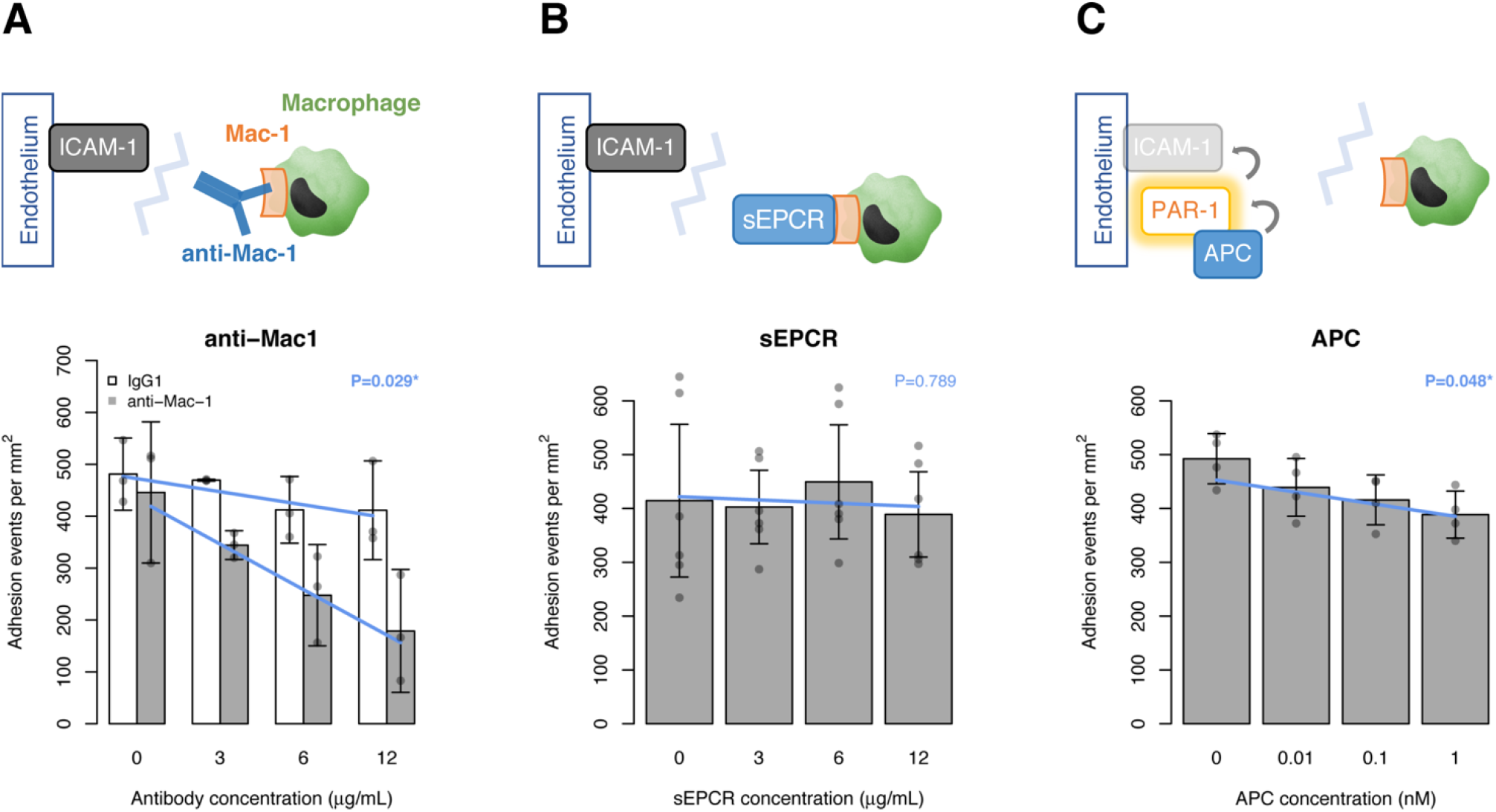
Leukocyte–endothelial cell adhesion in the presence of anti-Mac-1 antibody, sEPCR and APC. We performed static adhesion assays using PMA-stimulated monocytic cells (U937) and TNF-α-activated endothelial cells (HUVEC). We quantified the leukocyte–endothelial cell adhesion events after incubation with increasing concentrations of anti-Mac-1 antibody and matched IgG control, as well as recombinant sEPCR and APC, in each separate wells of the assay (**Methods**). Using a phase-contrast video-microscope, 5-sec videos were recorded at five different fields at random, and cell adhesion events (i.e. rolling, arresting and transmigration) quantified. Blue lines indicate the fitted linear regression models. Error bars show standard errors of the means. All experiments were performed in triplicate. <u>Credits:</u> The immune response, Big Picture (http://bigpictureeducation.com/).

## Discussion

Elucidation of the molecular basis of cross-disease associations affords a major opportunity to advance understanding of disease etiology. Leveraging recent advances in population biobanks, statistical genomics and translational epidemiology, we illustrate an integrative, multi-modal approach to address this challenge. We applied this approach to two vascular diseases oppositely associated with the missense variant p.S219G (rs867186) in *PROCR*. We showed that *PROCR*-219Gly protects against CAD but increases susceptibility to VTE through distinct chains of molecular events, summarized in **Fig. 7**.

**Figure 7.**
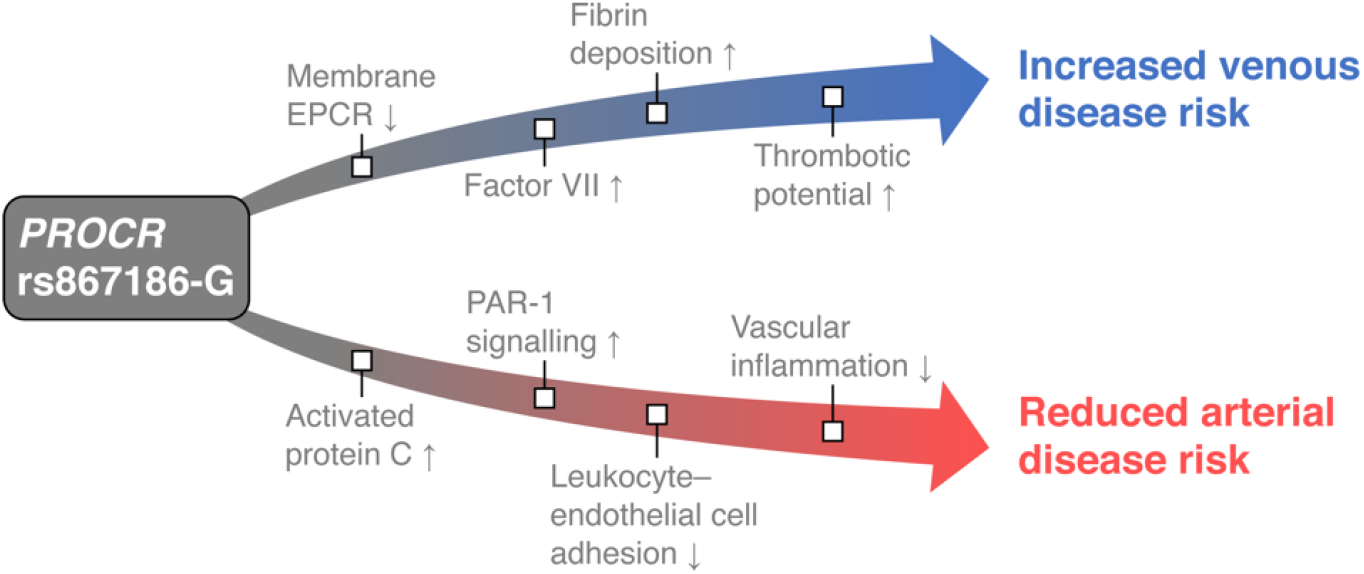
Proposed molecular chain of events underlying the PROCR-p.S219G variant.

Several aspects of our approach are generalizable to the study of other cross-disease associations (**Fig. 1**). First, the availability of large, disease-agnostic population biobanks with linked genomic, molecular phenotype and health record data, such as UK Biobank, provides an opportunity to systematically characterize the molecular underpinnings of health outcomes. Second, publicly available bioinformatics tools, including SAIGE ^29^ and PhenoScanner ^57^, allow for the mining of these data and the generation of specific hypotheses about the underlying biological mechanisms at individual genetic association loci. Third, the use of freely available software for statistical colocalization ^44^ and Mendelian randomization ^58^ analyses enables evaluation of the extent to which associated phenotypes share the same causal variant and the causal relationship of molecular biomarkers with a disease outcome. Fourth, the increasing availability of participants in bioresources (e.g. NIHR BioResource) who have agreed to invitation to biomedical studies on the basis of their genetic and/or phenotypic characteristics enables targeted mechanistic studies tailored to specific hypotheses. This includes recall-by-genotype studies, which afford an efficient approach to detailed phenotyping that can be applied to different study designs, biological samples and experimental techniques ^46^.

The data from our study show that *PROCR*-219Gly leads to a perturbed PC pathway, which acts focally to modulate the circulating levels of APC and has downstream effects on the biological mechanisms of associations with VTE and CAD. In our phenome-scan, we found a significant association between *PROCR*-219Gly and higher plasma levels of APC (**Fig. 1**). APC is a well-known anticoagulant that has also been shown to function as a cytoprotective and anti-inflammatory agent via the protease-activated receptor 1 (PAR-1) ^59^. Indeed, endothelial cells exposed to APC have reduced surface levels of key intercellular adhesion molecules, such as intercellular adhesion molecule-1 (ICAM-1) and vascular cell adhesion molecule-1 (VCAM-1), which regulate the adhesion of leukocytes to the endothelium ^60-62^. By performing static adhesion assays, we provided evidence that increasing concentrations of APC reduce the adhesion of activated monocytes to endothelial cells (**Fig. 6**). Based on these data, we propose that the APC/PAR-1 signaling pathway may be critical to protecting against CAD by reducing leukocyte–endothelium adhesion and vascular inflammation in carriers of *PROCR*-219Gly.

Our findings have implications for therapeutic strategies targeting the PC pathway for vascular diseases. Despite early positive clinical data that proposed the use of Drotrecogin alfa (Xigris^®^; a recombinant form of APC) as a therapeutic intervention for sepsis and septic shock, the medicine was withdrawn due to the lack of replication in subsequent trials and its associated risk of bleeding ^63^. APC has since emerged as a potential candidate for the treatment of stroke. Clinical trials are currently ongoing to test in patients with acute ischemic stroke the efficacy of 3K3A-APC, a recombinant form of APC that lacks its anticoagulant activity but retains its PAR-1 cell-signaling activities ^64^. Preliminary results showed that patients receiving 3K3A-APC had reduced hemorrhage volume and hemorrhage incidence on day 30 following initial drug infusion, relative to a placebo group ^65^. The findings from these clinical studies are consistent with the results of our wide-angled genetic association scan (**Fig. 2**), and provide a rationale to define and catalogue the disease relationships of pleiotropic variants on a genome-wide level to inform the development of new medicines.

Our phenome-scan also showed a significant association of *PROCR*-219Gly with higher plasma levels of FVII (**Fig. 2**). Recently, FVII has been identified as a ligand for EPCR and shown to bind EPCR with the same affinity as PC ^66^. Although EPCR does not affect the activation of FVII, the interaction of EPCR with FVII leads to the clearance of FVII/FVIIa from the circulation through endocytosis ^66^. Our data are consistent with this observation, as *PROCR*-219Gly is not only associated with higher plasma levels of FVII but also increased shedding of the membrane-bound EPCR (**Fig. 5**). Thus, the reduced availability of EPCR could directly contribute to the reduced internalization of FVII/FVIIa and increased accumulation in the circulation, which in turn may increase thrombotic potential. This proposed mechanism is consistent with the suggestive signal observed in *PROCR*-219Gly carriers for higher levels of D-dimer ^36^, a marker of blood clot degradation, as well as the genome-wide significant association observed with shorter prothrombin time ^13^. Nonetheless, further studies are needed to elucidate the complex interaction between EPCR and its ligands PC (APC) and FVII (FVIIa), as well as the downstream consequences of these interactions on hemostasis.

Taken together, our study provides new insights into the role of the PC pathway in arterial and venous diseases. We demonstrate that the combination of population biobank data and advanced statistical methods can help identify causal biomarkers and pathways, and that recall-by-genotype is a powerful experimental approach that can yield informative mechanistic insights. Overall, our study provides a framework for mapping molecular mechanisms that underlie cross-phenotype associations.

## Material and Methods

### PROCR-rs867186 phenome-scan

To assess the effects of *PROCR*-rs867186 genotype on cardiovascular intermediate traits and outcomes, we collated data from the latest available genome-wide association studies using PhenoScanner v2, a database of human genotype-phenotype associations ^57^. To allow comparative analyses, we only considered data from individuals of European ancestry. We focused our analyses on cardiometabolic traits and outcomes; thus, not all genome-wide significant associations are reported. **Suppl. Table 1** provides an overview of all data used in our analyses. The results of the complete data query can be found at: http://www.phenoscanner.medschl.cam.ac.uk/?query=rs867186&catalogue=GWAS&p=5e-8&proxies=None&r2=0.8&build=37.

### Determination of equilibrium binding constants

Equilibrium binding constants (K_d_ values) of modified aptamers were determined by filter binding assay. K_d_ values of modified aptamers were measured in SB18T buffer (40 mM Hepes pH 7.5, 102 mM NaCl, 5 mM KCl, 5 mM MgCl_2_, 0.01% Tween-20). Modified aptamers were 5’ end-labeled using T4 polynucleotide kinase (New England Biolabs) and γ-[^32^P]ATP (Perkin-Elmer). Commercially available proteins to be used in the filter binding assay (protein C, APC, sEPCR, thrombin, factor V, factor VIIa, protein S and thrombomodulin) were biotinylated by covalent coupling of EZ-Link NHS-PEG4 -Biotin (Thermo Scientific) following the manufacturer’s protocol. Briefly, proteins were combined with a 10-fold molar excess of EZ-Link NHS-PEG4 -Biotin in SB18T buffer and incubated at room temperature for 30 min. Free biotin was removed via YM-3 filtration (Millipore). Following biotinylation, protein concentrations were determined using a Micro BCA Protein Assay kit (Thermo Fisher). Radiolabeled aptamers (∼20,000 CPM, 0.03 nM) were mixed with biotinylated proteins at concentrations ranging from 10^−7^ to 10^−12^ M and incubated at 37°C for 40 min. Bound complexes were partitioned on MyOne streptavidin beads (Invitrogen) and captured on Durapore filter plates (EMD Millipore) and the fraction of bound aptamer was quantified with a phosphorimager (Typhoon FLA 9500, GE) and data were analyzed in ImageQuant (GE). To determine binding affinity, data were fit using the equation: y = (max – min)(Protein)/(K_d_ + Protein) + min.

### Competition binding assays

Competition binding assays were performed to test whether sEPCR and thrombomodulin interfere with the SOMAmer reagent 2961-1_2 binding to protein C or whether sEPCR, protein S and factor VIIa interfere with the binding of SOMAmer reagents 2961-1_2, 3758-63_3 and 3758-68_3 to APC. These experiments were performed by pre-incubating equal volumes of biotinylated protein C (80 nM) or biotinylated APC (48 nM or 80 nM) with competitor protein concentrations ranging from 10^−5^ to 10^−10^ M at 37°C for 30 min in SB18T buffer in the presence of 2 µM polyanionic competitor Z-block (a 30-mer modified DNA sequence, [AC(BndU)_2_]_7_AC)^23^ to allow protein complexes to form. Following the 30-min incubation, the reaction was diluted in half with radiolabeled SOMAmer reagent (20,000–60,000 CPM, 0.03 nM) and returned to 37°C for an additional 30 min. Bound complexes were partitioned on MyOne streptavidin beads and captured on Durapore filter plates and the amount of bound aptamer was quantified with a phosphorimager and data were analyzed in ImageQuant. The fraction of SOMAmer bound at each competitor concentration was normalized to the signal in the no competitor control well.

### Multi-trait colocalization

We performed colocalization analysis at the *PROCR* gene locus (chr20: 31,916,110–35,505,723 bp; hg19), as defined by linkage disequilibrium (LD) (r^2^ <0.2 to rs867186; based on the 1000 Genomes Project data). Details about the GWAS summary statistics used for this analysis are provided in **Suppl. Table 1**. Variants with both imputation (INFO)-score <0.7 and MAF <0.01, or variants with INFO-score <0.3 and MAF >0.01 were removed. The remaining 4,264 SNPs shared across each of the datasets were aligned to the DNA plus-strand (hg19) prior to colocalization analyses. We used a Bayesian algorithm, implemented in the Hypothesis Prioritization in multi-trait Colocalization (HyPrColoc) method ^44^, to perform colocalization across all traits simultaneously. HyPrColoc extends the established coloc methodology ^67^ by approximating the true posterior probability of colocalization with the posterior probability of colocalization at a single causal variant and a small number of related hypotheses ^44^. If all traits do not share a causal variant, HyPrColoc employs a novel branch-and-bound selection algorithm to identify subsets of traits that colocalize at distinct causal variants at the locus. We used uniform priors as primary analysis, which used strong bounds for the regional and alignment probabilities as default, i.e. the PR* (regional probability threshold) = PA* (alignment probability threshold) = 0.7, so that the algorithm identified a cluster of colocalized traits only if PRPA >0.49. We also performed sensitivity analysis with non-uniform priors to access the choice of priors, which used a conservative trait-level prior structure with *P*=1×10^−4^ (prior probability of a SNP being associated with one trait) and 1–γ=0.02 (1–γ is the prior probability of a SNP being associated with an additional trait given that the SNP is associated with at least one other trait), i.e. 1 in 500,000 variants is expected to be causal for two traits.

### Selection of instrumental variables for MR analysis

We obtained regional association statistics at the *PROCR* region for plasma PC levels from the ARIC study and plasma APC levels from the INTERVAL study to assess the causal effects of PC (APC) on cardiovascular outcomes. Details about the GWAS data on cardiovascular outcomes are provided in **Suppl. Table 1**. To select genetic variants as instrumental variables for PC levels, we first removed SNPs with MAF <0.01 and INFO-score <0.8. Next, we performed LD clumping to obtain approximately independent SNPs. In brief, the algorithm groups SNPs in LD (r^2^ ≥0.1 in 4,994 participants from the INTERVAL study ^68^) within ±1 MB of an index SNP (i.e. SNPs with association *P*-value ≤5×10^−8^). The algorithm tests all index SNPs, beginning with the smallest *P*-value and only allowing each SNP to appear in one clump. Thus, the final output contains the most significant protein-associated SNPs for each LD-based clump across the genomic region. An overview of the instrumental variables is provided in **Suppl. Table 3**. This analysis was performed using PLINK v1.90 ^69^.

### Mendelian Randomization analyses

We used two-sample Mendelian randomization (MR) ^58,70^ to estimate the causal associations between PC and cardiovascular outcomes. The MR approach was based on the following assumptions: (i) the genetic variants used as instrumental variables are associated with PC levels; (ii) the genetic variants are not associated with any confounders of the exposure-outcome relationship; and (iii) the genetic variants are associated with the outcome only through changes in PC levels, i.e. a lack of horizontal pleiotropy. We applied the inverse-variance weighting (IVW) method in a multiplicative random-effect meta-analysis framework ^70^, MR-Egger regression ^71^ and MR-PRESSO ^72^ to estimate the causal effects. We also performed several sensitivity analyses to assess the robustness of our results to potential violations of the MR assumptions, given these analyses have different assumptions for validity: (i) heterogeneity was estimated using the MR-IVW *Q*-statistic; (ii) horizontal pleiotropy was estimated using MR-Egger’s intercept; (iii) influential outlier instrumental variables due to pleiotropy were identified using MR-PRESSO; and (iv) MR-Steiger filtering ^73^ was used to eliminate spurious results due to reverse causation. We also applied reverse MR ^74^ to evaluate evidence for causal effects in the reverse direction by modelling disease phenotypes as the exposure and PC level as the outcome. Instrumental variants for phenotypes of interest (i.e. CAD, DVT/VTE) were selected from their original GWAS data (**Suppl. Table 1**). The effects of these GWAS SNPs on PC levels were derived from Sun et al ^31^. The power and strength of the instrumental variables was assessed using the variance explained (*R*^*2*^) and F-statistics ^75^. The MR analyses were conducted using the MendelianRandomization ^58^, TwoSampleMR ^76^ and MR-PRESSO ^72^ packages in R v3.4.2.

### Recall-by-genotype study

The study was approved by the Leicester Central Research Ethics Committee and Health Research Authority (Reference: 17/EM/0028). Healthy volunteers were recruited from the NIHR Cambridge BioResource with informed consent. Participants who were older than 18 years of age and of European ancestry were selected based on *PROCR*-rs867186 genotype and homozygosity of the major allele for both *F5*-R506Q (rs6025; Factor V Leiden) and *F2*-G20210A (rs1799963; Factor II). Participants across the three rs867186 genotype groups were matched at the end of the study with respect to sex and age (within 10 years). Study participants were excluded that had a diagnosis of (i) a chronic disease; (ii) hypertension (or history of consistently high blood pressure readings, i.e. >140/90 mmHg); and/or (iii) hypercholesterolemia (or history of consistently high cholesterol levels, i.e. >6 mmol/l). Participants agreed to fast and abstain from caffeinated drinks for at least four hours prior to the study visit and to not receive any vasoactive medication for up to seven days prior to procedures.

### Assessment of baseline characteristics of study participants

Participants reported past medical conditions, demographic factors (e.g. ethnicity) and lifestyle factors (e.g. smoking and alcohol consumption). Height and weight/body fat were measured using a stadiometer and bioelectrical impedance (i.e. Tanita scale), respectively. Blood pressure and heart rate were assessed in 1-min intervals using a validated, automated device while seated and again after 3–5 min standing. All measurements were done in triplicate using the same arm. An overview of the characteristics of the study participants is provided in **Suppl. Table 5**. These characteristics are presented as mean and standard deviation or percentage. Continuous and categorical variables between homozygous groups were compared using the 2-sample t-test and chi-square test, respectively.

### Blood sample collection and processing

A total of 46 ml of peripheral blood was collected from each donor using a 21 gauge needle unless clinically contraindicated. We collected blood in two S-Monovette 7.5-ml K3 EDTA tubes and two S-Monovette 10-ml sodium citrate 3.2% (1:10) 9NC tubes (Sarstedt). Samples were immediately centrifuged at 4°C and 1,000×g for 15 min. Multiple aliquots of the top phases were stored at - 80°C within 30 min of blood draw. In addition, we collected blood in two S-Monovette 4.9-ml serum Z-Gel tubes (Sarstedt). These samples were centrifuged at room temperature and 2,500×g for 10 min, and then immediately stored at -80°C until use. A full blood count for all donors was obtained from blood collected in a S-Monovette 1.2-ml K3 EDTA tube using a Sysmex Hematological analyzer.

### Quantification of plasma biomarkers

Samples were thawed at 37°C for 15 min, mixed and then centrifuged at room temperature and 3,000×g for 10 min immediately prior to assay. Soluble EPCR levels were determined using an Asserachrom sEPCR kit (00264; Diagnostica Stago); thrombin/antithrombin III complex levels using an Enzygnost TAT micro immunoassay (OWMG15; Siemens Healthcare Diagnostics Limited); APC levels using an Activated Protein C assay kit (CSB-E09909H; Cusabio Biotech); and PC levels using a HemosIL Protein C chromogenic assay (0020300500; Instrumentation Laboratory). All assays were performed according to the manufacturer’s instructions. Samples were analyzed in random order and laboratory staff were blinded to genotype status. Participants with biomarker levels (or activity levels) 3 standard deviations above or below the population mean were removed.

### Tissue culture

All cultures were maintained at 37°C in a humidified chamber at 5% CO_2_. Human Umbilical Vein Endothelial Cells (HUVECs) (PromoCell) were cultured using an Endothelial Cell Growth Media (EGM)-Plus BulletKit (Lonza). U937 cells (ATCC) were suspended in RPMI-1640 Medium with GlutaMAX supplement, 10% fetal bovine serum (FBS), 100 U/ml penicillin and 100 U/ml streptomycin (ThermoFisher). Cells were used in experiments at passages 2–4.

### In vitro static adhesion assay

To quantify U937–HUVEC interactions, we used an *in vitro* static adhesion assay, as previously described ^77,78^. U937 cells were seeded at a density of 1×10^6^ in T25 flasks and differentiated into macrophages in the presence of 100 ng/mL phorbol 12-myristate 13-acetate (PMA) for 48 hours. HUVECs were seeded 24 hours prior at a density of 1×10^5^ cells in wells of a 12-well plate, and then treated with 10 ng/mL TNF for 4 hours on the day of the assay. Immediately before commencing the assay, U937 cells were collected and re-suspended in fresh medium at a concentration of 1×10^5^ cells/ml. HUVEC monolayers (at ≥90% confluence) were rinsed in Phosphate NaCl (PBSA) buffer and incubated with 1 ml U937 cell (i.e. 1×10^5^ cells) suspension for 5 min at 37°C. After aspirating the U937 suspension, the HUVECs and any adherent U937 cells were gently rinsed four times in PBSA, and then a further 2 ml PBSA were added to the well. Using a phase-contrast video-microscope (Leica Microsystems, DMI3000B), 5-sec video recordings at 20-fold magnification were made, choosing five different fields at random. Quantification of cell adhesion events (i.e. rolling, arresting and transmigration) was performed using the ImagePro v6.3 software.

### Antibodies

Allophycocyanin (APC)-conjugated Rat Anti-Human EPCR monoclonal antibodies derived from two different clones were obtained from BD Biosciences (563622; clone: RCR-252) and Thermo Fisher Scientific (17-2018-42; clone: RCR-227). Corresponding APC-conjugated Rat IgG1, κ Isotype Control antibodies were sourced from BD Biosciences (554686; clone: R3-34) and Thermo Fisher Scientific (17-4301-82; clone: eBRG1). An unconjugated Rat Anti-Human EPCR monoclonal antibody was obtained from BD Biosciences (552500; clone: RCR-252).

### Quantification of membrane EPCR levels by flow cytometry

We first confirmed the suitability of allophycocyanin (APC)-monoclonal antibodies to EPCR from two distinct clones for flow cytometry using HUVECs as a positive control cell line. We lysed 100-μl citrated whole blood samples for 10 min at room temperature using Lysing Solution 10X Concentrate (349202; BD Biosciences). Lysed blood was then centrifuged at 600×g for 6 min and 4°C, and the pellet re-suspended in HEPES buffered saline (Sigma-Aldrich). Cultured cells were also re-suspended in HEPES buffered saline, to a final concentration of 10^5^ cells/100 μl. Rat Anti-Human EPCR monoclonal antibodies and isotype controls were added as appropriate at a final concentration of 0.125 μg/100 μl and incubated for 20 min at room temperature in the dark. Samples were diluted in 0.5 ml ice-cold HEPES buffered saline prior to flow cytometric analysis using either a Cytomics FC500 or a CytoFLEX S Flow Cytometer (Beckman Coulter). CD14^+^ Monocytes and CD16^+^ neutrophils from blood lysates were gated using forward and side light scatter, enabling discrimination by cell size and granularity, respectively. Results were recorded as median fluorescence intensity (MFI).

## Data Availability

Data available on request from the authors.

## Acknowledgements

We gratefully acknowledge the participation of all National Institute for Health Research (NIHR) BioResource Centre Cambridge volunteers, and thank the NIHR BioResource Centre Cambridge and staff for their contribution. We thank the NIHR and NHS Blood and Transplant. This work was supported by the British Heart Foundation Cambridge Centre of Excellence [RE/13/6/30180]. The Cardiovascular Epidemiology Unit is supported by core funding from the: UK Medical Research Council [MR/L003120/1], British Heart Foundation [RG/13/13/30194; RG/18/13/33946] and NIHR [Cambridge Biomedical Research Centre at the Cambridge University Hospitals NHS Foundation Trust]. This work was further supported by Health Data Research UK, which is funded by the UK Medical Research Council, Engineering and Physical Sciences Research Council, Economic and Social Research Council, Department of Health and Social Care (England), Chief Scientist Office of the Scottish Government Health and Social Care Directorates, Health and Social Care Research and Development Division (Welsh Government), Public Health Agency (Northern Ireland), British Heart Foundation and Wellcome. J.E.P. is supported by a UKRI Innovation Fellowship at Health Data Research UK (MR/S004068/1). J.D. is funded by a NIHR Senior Investigator Award. J.M.M.H. and A.M. are funded by the NIHR [Cambridge Biomedical Research Centre at the Cambridge University Hospitals NHS Foundation Trust]. The views expressed are those of the authors and not necessarily those of the NHS, the NIHR or the Department of Health and Social Care. The US National Heart, Lung and Blood Institute (NHLBI) provided support for LITE via R01HL059367. This work was carried out in part using computing resources at the University of Minnesota Supercomputing Institute. The Atherosclerosis Risk in Communities (ARIC) study has been funded in whole or in part with US federal funds from the National Heart, Lung and Blood Institute (HHSN268201700001I, HHSN268201700002I, HHSN268201700003I, HHSN268201700004I, HHSN268201700005I, R01HL087641, R01HL086694); National Human Genome Research Institute (U01HG004402); and National Institutes of Health (HHSN268200625226C). The authors thank the staff and participants of the ARIC study for their important contributions. Infrastructure was partly supported by Grant Number UL1RR025005, a component of the National Institutes of Health and NIH Roadmap for Medical Research. M.S-L is a recipient of a *Miguel Servet* contract from the Spanish Ministry of Health (ISCIII CP17/00142). P.S. de Vries is supported by American Heart Association grant number 18CDA34110116. The CHARGE Hemostasis Working Group acknowledges a NIH R01 HL134894 grant. SomaScan^®^ and SOMAmer^®^ reagent are registered trademarks of SomaLogic, Inc. This research has been conducted using the UK Biobank resource under Application Number 26865.

